# Using outlier detection methods to incorporate highly heterogeneous infection rates into compartment models

**DOI:** 10.64898/2026.06.22.26355953

**Authors:** Lennart Schüler, Peter Lünenschloß, David Schäfer, Jan Bumberger, Justin M. Calabrese

## Abstract

Superspreading events (SSEs) produce extreme, rare bursts of disease transmission that standard compartment models, which assume population homogeneity, fail to capture. This inability to model heterogeneity in transmission rates can result in biased estimates of transmissivity. To address this limitation, we present a modular framework that treats SSEs as statistical outliers in case count time series and incorporates them into SIR-type models via pulse terms that transfer SSE cases directly from susceptible to infected compartments. This separation isolates anomalous SSE-driven transmission from background spread, which reduces bias when estimating mean transmission rates. We validate the approach on synthetic data generated by a stochastic model with embedded SSEs, demonstrating accurate recovery of the true non-SSE transmission parameter. We then apply the method to COVID-19 outbreaks in Hong Kong and the German district of Gütersloh, showing improved model fits and more robust estimates of background transmissivity both for a period with constant transmission and for a period with temporally structured NPI-driven heterogeneities. The framework’s interchangeable outlier-detection, compartment, and SSE modules make it adaptable to diverse diseases and data contexts.

## Introduction

Superspreading events (SSEs), which occur when a case causes more infections than would be expected in 99% of cases in a homogeneous population [1], are known to strongly influence the dynamics of a wide range of diseases. The SARS outbreak of 2002 – 2004 provides a particularly well-studied example [1–7], where most of the infected population hardly contributed to the epidemic, with over 73% of the cases having an individual reproduction number of less than one and a small proportion mainly driving disease spread, with 8% having an individual reproduction number of over 8 [1]. Other diseases where influential SSEs have been identified include Measles, Influenza, Rubella, Smallpox, Ebola Haemorrhagic Fever, and SARS-CoV-2 [1, 8–14]. In the context of COVID-19, SSEs typically take place in settings where many people are in close proximity over an extended period of time. Examples for such settings include cruise ships [9], churches [10], care homes [11], homeless shelters [12], prisons [13], conferences [8], and factories where employees work in close proximity [14]. SSEs occurring under such conditions result in extremely skewed transmissivity distributions, which imply highly heterogeneous transmission rates. In other words, SSEs create a situation in which the mean transmission rate captures neither the “typical” disease dynamics nor the anomalous transmission that occurs during (and immediately after) an SSE.

Compartment disease models have long formed the backbone of mathematical epidemiology [15–20]. Variations of these models have been applied to outbreaks spanning a broad array of diseases [21–27], and are often used to inform disease mitigation strategies and guide public health policies [25, 26, 28–35]. In particular, compartment models featured prominently in a huge number of both theoretical and applied studies examining various aspects of the COVID-19 pandemic (see Brauer et al. 2019 [36] and examples therein). However, despite their ubiquity and utility, compartment models make extremely strong assumptions on population homogeneity. Every individual is placed into one of several compartments, within which they all have exactly the same transition rates such as latent period, recovery time, or transmissivity. Of these, transmissivity stands out as being particularly important, because it controls a super-linear term in compartment models and thus has an outsized influence on overall disease dynamics.

The extreme heterogeneity in transmissivity that SSEs engender is thus not well-captured by standard compartment type models. While the homogeneity assumptions underpinning compartment models have frequently been criticized [6, 32, 37, 38], these models continue to be used in situations where SSEs are likely to be important. In particular, the COVID-19 pandemic has highlighted the inability of compartment models to account for highly variable transmission rates, which occur especially during SSEs [39, 40]. In a previous work we took a first step toward integrating heterogeneities in transmission rates into SIR-type compartment models [40]. The focus of that study was to model the transmission dynamics resulting from changing non-pharmaceutical interventions (NPIs), which are predictable, temporally structured heterogeneities shaped by policy decisions and behavioural changes [41]. The NPI induced changes in transmission rates were implemented by piecewise constant parameters, striking a balance between meaningful parameterisation and robust fitting. In contrast to models with daily varying parameters, our approach preserves the simplicity of the original SIR model, which has constant parameters, while still allowing for a more nuanced modelling of transmission dynamics. In this paper, we revisit this model and expand to include SSEs, which introduce a distinct form of heterogeneity characterised by their unpredictability and potential to create extreme variations in transmission rates.

Superspreading events have been defined in statistical terms [1]. This definition is highly analogous to the concept of outlying data points in univariate time series. For example, the Z-score outlier detection method defines outliers by using a specific percentile of the underlying distribution (e.g. [42]). A further correspondence between the two concepts is that statistical outliers often arise from anomalous events, and SSEs are, by definition, anomalous events with respect to the typical transmission dynamics of a focal disease. Outlier detection in time series data is a very well-studied topic in statistics due to its relevance in many different scientific domains [42–45]. Furthermore, many outlier detection algorithms feature robust and widely-used implementations in popular data-science-oriented programming languages such as Python and R.

Here, we leverage the analogy between the definition of SSEs in epidemiology and that of outlying data points in statistics to introduce an innovative modular method for incorporating highly heterogeneous transmission rates into the compartment modeling framework. Our approach flexibly combines interchangeable statistical outlier detection methods with modified compartment models. In particular, observations flagged as outliers in a case count time series are transferred from the susceptible to the infected compartment via pulse terms on the right-hand side of the compartment model equations. This creates a different channel through which susceptible individuals are infected during an SSE, and effectively separates SSEs from the typical transmission dynamics, which we call background transmission rates throughout this paper.

By doing so, the mean transmission rates of compartment models can be estimated with less bias in the presence of SSEs because they no longer have to account for the skewness induced by anomalous transmission events. The framework’s modular design makes it highly flexible. The outlier detection method, compartment model, and SSE model can be replaced individually with alternatives that are more appropriate for a specific problem. We demonstrate our approach with the Local Outlier Factor (LOF) method, which is an easy to understand and readily available outlier detection method with interpretable parameters.

We first validate our approach via a generative stochastic model that creates synthetic datasets where the background infection curve created by a standard SIR-type model is punctuated with occasional SSEs. This allows us to compare the estimate of the background disease transmission rate obtained by our approach in the presence of stochastic SSEs against the known true background transmission value used in the generative model.

We then demonstrate the real-world performance of our method on COVID-19 spread in two different regions. Specifically, we apply our method to Hong Kong (pop. 7,500,000) and the German district of Gütersloh (pop. 360,000). For Hong Kong, we chose a time period in which the transmission rate can be modelled as a constant to first demonstrate the results of including SSEs into classical SIR models. For Gütersloh, we employ the Schüler et al. 2021 model to simulate the complete first wave and the subsequent summer characterised by mostly low case numbers. Here, the temporally structured heterogeneities through NPI changes play an important role and we demonstrate the results of including SSEs as a different kind of heterogeneity. In both cases, the model fits improve, resulting in a better estimation of the background transmission rates.

## Materials and methods

### Modified SSE-SIR type compartment models

We begin with the SIR model introduced by Schüler et al. 2021 [40] and successfully applied at multiple levels of data resolution in Germany. The advantage of this model is that it accounts for the heterogeneities induced by NPI changes by allowing the transmission rate to change in a step-wise manner as NPIs are applied or lifted. If no NPI changes occur, the model is identical to the classical SIR model. The model is expressed through a system of 3 coupled ordinary differential equations, one for each of the considered compartments *S*usceptible, *I* nfectious, *R*ecovered:

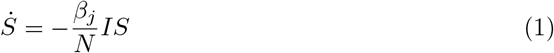

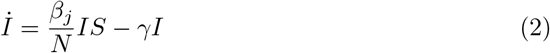

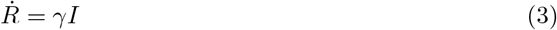

with *N* being the total population, the contact rates *β*_*j*_ creating a step function, which only changes when non-pharmaceutical interventions (NPIs) are introduced or lifted and a constant recovery rate *γ*. However, this approach cannot adequately model SSEs [40]. The reason for this limitation is the underlying assumption that the whole population has homogeneous transmissivity within each time partition defined by NPI change.

To take SSEs or other strongly inhomogeneous effects into account without altering the otherwise successful structure of (1) -(3), we use an outlier detection method, which will be introduced later, to identify these anomlies. This is done as a preprocessing step before the actual parameter inference or simulation. The detected outliers are then used to create a second infection channel transferring individuals from the *S* to the *R* compartments.

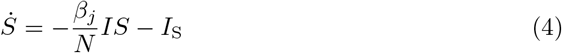

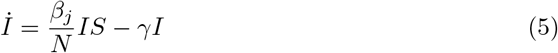

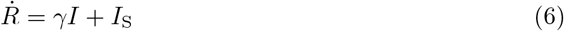

with *I*_S_ being the residual cases per day, in contrast to *I*, which is the current number of infectious individuals.

For modelling the transmissions of the SSE infection channel, we assume that the number of transmissions during an SSE suddenly jumps to its maximum value *a* and then decays according to the Weibull function for the duration of the SSE *t*_S_ to a final value given by *τ*.

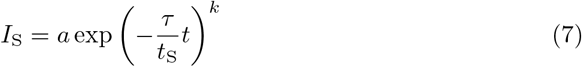

We found that the Weibull function, as a generalisation of the exponential function, strikes a good balance between good fits and too many parameters. The residuals of the detected outliers from the preprocessing step are calculated by fitting the SIR model (1) - (3) to the reported cases without the outliers and then subtracting the modelled cases from the outliers. These residuals are used to parametrise the SSE model. Specifically, we need the duration of an SSE *t*_S_ and its maximum value or amplitude *a*. In case data in between detected SSEs are not flagged as outliers, we additionally flag all data in gaps between detected SSEs with a maximum gap of 3 days. The SSE duration *t*_S_ is then set top the total duration of the identified SSE. The amplitude *a* is set to the maximum detected value of the SSE. The final two parameters *θ* and *k* are estimated via non-linear least squares.

### Generative model and simulation study

A generative model of combined SIR-SSE dynamics allows us to create synthetic data with which we can test the performance of the modified SIR-SSE model (4) -(6). We assume the SSEs to be independent of each other and thus we model their timing as a Poisson point process (PPP)

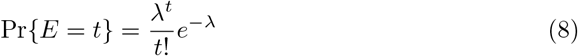

with *E* being the random occurrence of an SSE on day *t* and *λ* being the rate of the PPP. The amplitude parameter *a* of the SSE model (7) is drawn from a Lognormal distribution, the duration *t*_S_ from a discrete uniform distribution and the parameters *τ* and *k* both from a truncated normal distribution, making the generative model a compound PPP. It should be noted that the PPP in this form only works for *t* ∈ N, which would only be violated if we would be interested in a sub-daily temporal resolution of the events.

In order to perform a systematic model intercomparison between the SIR model (1) -(3) and the SSE-SIR model (4) -(6), we set up the generative model described above, by using the NPI change points from Germany [40]. We took the estimated transmission rates 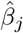 from Gütersloh without the detected SSE as the mean values of truncated normal distributions 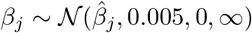 with a standard deviation of *σ* = 0.005 from which we drew the transmission rates for each time partition within a given realisation. We opted for such a small standard deviation, because the case numbers are extremely sensitive to changes in the transmission rates. For each realisation, we generated SSEs by sampling from the compound PPP (8) with a rate parameter of *λ* = 0.003. The amplitudes of the SSEs were drawn from *a* ∼ Lognormal(30, 0.5) and the maximum duration was sampled from *t*_S_ ∼ *U* (1, 16). The last two parameters were drawn from the truncated normal distributions *τ* ∼ *N* (4, 1 2, 4) and *k* ∼ *N* (0.95, 0.3 0.4, 1.5). These distributions were selected so that the Gütersloh SSE belongs to the largest possible outbreaks while preventing the generated SSEs from degenerating. For instance, small values of *τ* lead to rapidly decaying SSEs, rendering the duration parameter meaningless. Similarly, a small *k* value would result in minimal decay, negating the characteristic peak nature of SSEs. Finally, we added an error term to each data point drawn from Lognormal(ln *I*_gen_, 0, 2) for every time series, with *I*_gen_ being the generated time series.

We generated an ensemble of 1000 realisations from the compound PPP (8) to evaluate the performance of the fits of SSE-SIR model (4) - (6) with those of the classical SIR model (1) - (3). We utilised three key metrics for evaluation: the mean squared error (MSE, lower values are better) to assess accuracy, the symmetric signed percentage bias (SSPB, smaller absolute values are better) to measure bias, and the Pearson correlation coefficient (larger values are better) to evaluate the strength of association. The performance of the uLOF outlier detection method was evaluated with Youden’s J statistic [46, 47], also known as the true skill statistic.

### Local Outlier Factor outlier detection

The Local Outlier Factor (LOF) was first introduced by Breunig et al. (2000) [48] and has since been adapted many times, creating many different variants [45]. The popularity of the LOF derives from the method’s ease of use coupled with its readily estimable and straightforwardly interpretable parameters. Furthermore, the LOF is accessible via Python and R, making it an ideal first choice for outlier detection. We briefly summarize the method here, but more details can be found in the Appendix. The LOF assigns a score to data points in multi-dimensional datasets to indicate their anomaly status based on variations in local data densities, by evaluating the local neighbourhood of each point.

The implementation of the uLOF in the SaQC Python package [49, 50], we use for this study, is defined by the function signature:

`flagUniLOF(k=20, thresh=1.5, p=1, min_offset=0)`

Parameters include `k` (number of neighbours), `thresh` (cut-off threshold), `p` (norm type), and `min_offset` (to prevent overflagging or false positives, e.g. of small increases in low variance regimes or small context windows). This allows for the incorporation of domain knowledge into the anomaly detection process. The parameters can either be manually adjusted, with interpretable parameters proving to be especially helpful or they can be optimised.

### Datasets and empirical case studies

We applied the outlier detection and the SSE-SIR model (4) - (6) together with the SIR model (1) - (3)to time series of reported COVID-19 cases from the special administrative region of China Hong Kong, and from the district of Gütersloh in the German state North Rhine-Westphalia. We accessed the data through the Python interface pyCOCAP to a database in which we have merged several different data sources [51]. The data from Hong Kong was originally gathered by the John Hopkins University and the data from Gütersloh was published by the Robert Koch Institute (RKI).

Hong Kong, a densely populated region with approximately 7.5 million inhabitants, was one of the very few territories which followed a rigorous zero-COVID strategy that ultimately became unsustainable with the emergence of the Omicron variant. The NPIs to implement this strategy included border closures, mandatory hospitalisation for mild and even asymptomatic cases, and extended quarantine protocols. For this case study, we examined the aftermath of Hong Kong’s fourth wave. To give some context, a brief description follows: the wave began in mid-November 2020, characterised by a rapid surge in reported cases, peaking in December at 120 d^−1^, but quickly dropped to about a fourth by the beginning of January. However, before the end of the fourth wave, the numbers quickly rose again to values of around 80 d^−1^, resulting in a fifth, overlapping wave. Interestingly, very similar patterns occurred in Europe shortly after the Christmas holidays of 2020 [52]. The increased numbers were mainly due to an increase of local outbreaks in different areas and buildings of Hong Kong, leading to multiple lockdowns of up to 48 hours, affecting tens of thousands of residents. By mid-February new cases fell to about 15 d^−1^. In anticipation of the Spring Festival on 12 February, the authorities cautiously relaxed certain restrictions, permitting selective reopening of leisure venues and easing restaurant regulations. At the same time, the initial phase of vaccination deployment commenced by late February. During this phase of constant NPI measures, an SSE was identified in a fitness gym in March 2021 and a total of 102 cases could be linked to this single SSE [53] (See Fig. 1A).

**Fig 1.**
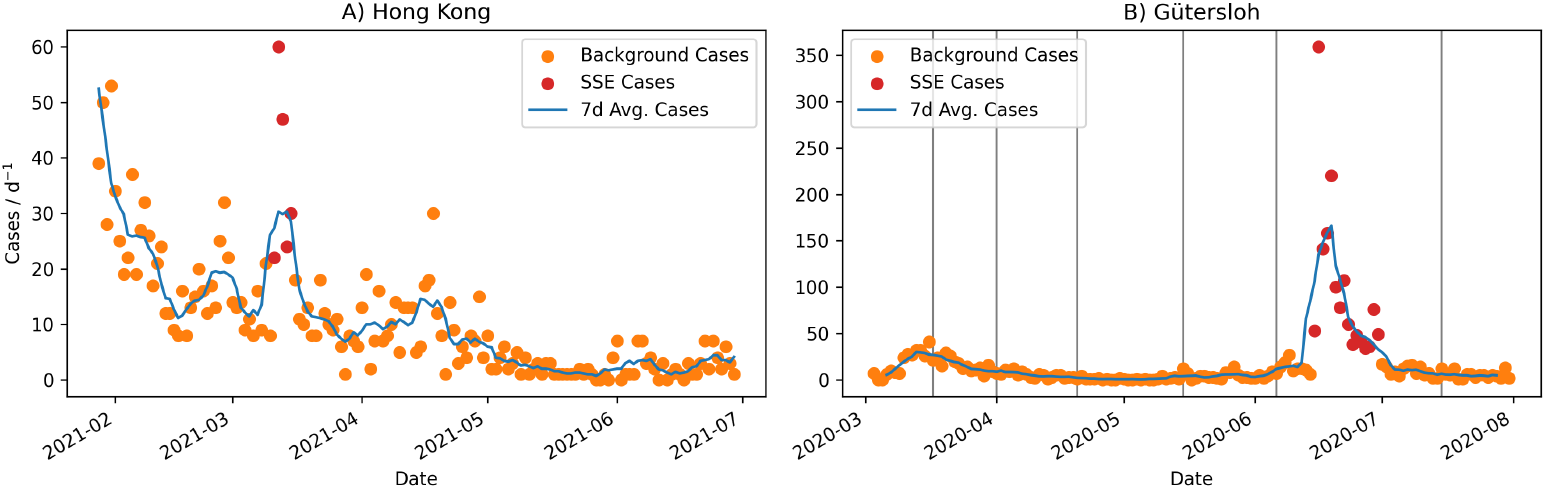
The reported daily COVID-19 cases (orange dots), together with the 7-day moving averaged cases (blue line) and the SSEs (red dots) for Hong Kong during its 4th wave (left panel) and for Gütersloh during its 1st wave (right panel). The black vertical lines indicate the dates of NPI changes in Germany.

The district of Gütersloh in North Rhine-Westphalia, Germany, home to approximately 360,000 residents, experienced a major SSE in June 2020 (see Fig. 1B) [14]. Prior to the SSE outbreak, Gütersloh experienced the COVID-19 pandemic much like many other districts in Germany, with its first cases reported in mid-March. During this period, strict NPI measures were implemented, including the closure of schools and kindergartens, the postponement of university semester starts, restrictions on visits to nursing homes, border closures, and even the enforcement of curfews. These measures effectively curtailed the first wave of the pandemic [31], ending it by April 2020. During the summer of 2020, Gütersloh experienced remarkably low infection rates, with reported cases between 0 and 4 on most days. In June 2020, the SSE outbreak occurred at the Tönnies slaughterhouse located in the city of Rheda-Wiedenbrück. On 17 June 2020, in response to the SSE, a complete production halt was imposed and all 7000 employees had to undergo a mandatory 14-day quarantine. After 16 days, the reported cases in Gütersloh had returned to pre-SSE levels. In total, 1413 employees were directly linked to the initial cluster, with an additional 353 secondary infections identified [54], making this SSE by far the largest one identified in Germany.

For Hong Kong, we concentrated only on the heterogeneity introduced by the SSE and excluded heterogeneity induced by NPI changes by selecting an interval around the SSE where the transmission rate did not change significantly, starting late January 2021 and ending early July 2021. Therefore, we fitted the SIR model (1) - (3) and the SSE-SIR model (4) - (6) to the reported case numbers with a single constant transmission rate *β*_1_. For Gütersloh, we extended the simulation time, starting at the beginning of the first wave on 3 March 2020 and and ending right before the start of the autumn wave on 1 August 2020. The dates of NPI changes were already identified in a previous work [40] and were used for the change points of the transmission rates *β*_*j*_.

## Results

### Model intercomparison

The results of the three performance metrics comparing the SIR and SSE-SIR model simulations (MSE, SSPB, and Pearson correlation coefficient) are presented as a violin plot (Fig. 2), showing the full distributions (blue and orange areas), the median (white line), the interquartile range (IQR, shown as a thick black bar), and the data that extends to 1.5 times the IQR (thin black bar) of the metrics. Our analysis revealed that the median performance as measured by all three metrics improved substantially when using the SSE-SIR model, as indicated by the white vertical lines in Fig. 2. This finding underscores a critical insight that even when outlier detection methods are not flawless, the approach can still lead to meaningful improvements in overall model performance. Notably, the Pearson correlation coefficient significantly improved, increasing from 0.66 to 0.96. Furthermore, each metric’s distribution for the standard SIR model, (1) - (3), is wider than the corresponding metric distribution of the SSE-SIR model. In particular, the extremely long tails of all three metric distributions of the standard SIR model were especially intriguing as they suggest that ignoring the heterogeneity induced by the SSEs will frequently lead to extremely biased and noisy results. The SSE model’s SSPB distribution is approximately symmetric, whereas the standard SIR model has a distribution characterized by the bulk of the probability density being symmetrically, though more broadly distributed and with a longer tail in the direction of overprediction. This suggests not only that the SSE-SIR model (4) - (6) outperforms the standard SIR model (1) - (3) in general, but also that there are critical scenarios where the latter completely fails to reproduce the underlying dynamics.

**Fig 2.**
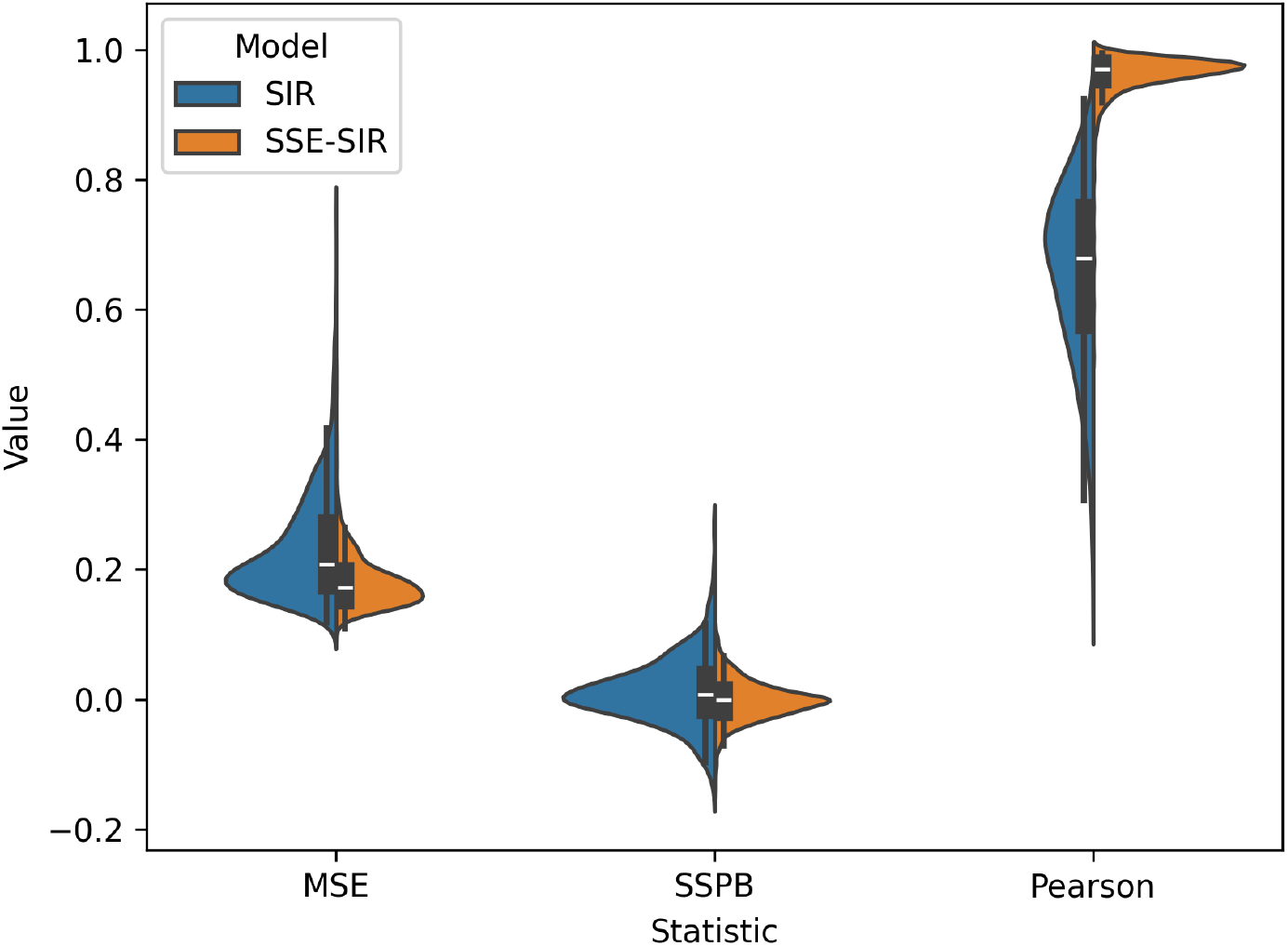
The mean squared errors (MSE), the symmetric signed percentage biases (SSPB), and the Pearson coefficients of 1000 model fits of the SIR model (1) - (3) (blue) and the SSE-SIR model (4) - (6) (orange) to the 1000 synthetically generated time series.

The performance of the uLOF outlier detection method is evaluated in the supplementary. We emphasise that the model performance improvements can be substantial, especially when looking at the long tails of the SIR model results, even though the outlier detection algorithm was chosen for its simplicity and interpretability, not because it showed the best performance among outlier detection methods for this particular application.

### Empirical case studies

We now examine two SSEs which were previously analysed in detail and for which we know the development over time. With this knowledge, we can model the SSEs with Eq. (7) and the temporal evolution of the daily reported case numbers with the SSE-SIR model (4) - (6), and compare its results to the standard SIR model (1) - (3).

The SSE in Hong Kong in March 2021 clearly stands out among the reported cases (Fig. 3) and the SSE model (7) describes this very narrow peak satisfactorily. Accounting for SSEs reduces the transmission rate from *β* = 0.124 d^−1^ to *β*_SSE_ = 0.109 d^−1^, a relative difference of 13%. When accounting for the SSE, the MSE improves from 1.1 to 0.6. The SSPB improves from -0.62 to 0.29, whereas the Pearson correlation coefficient does not change significantly from 0.72 to 0.74.

**Fig 3.**
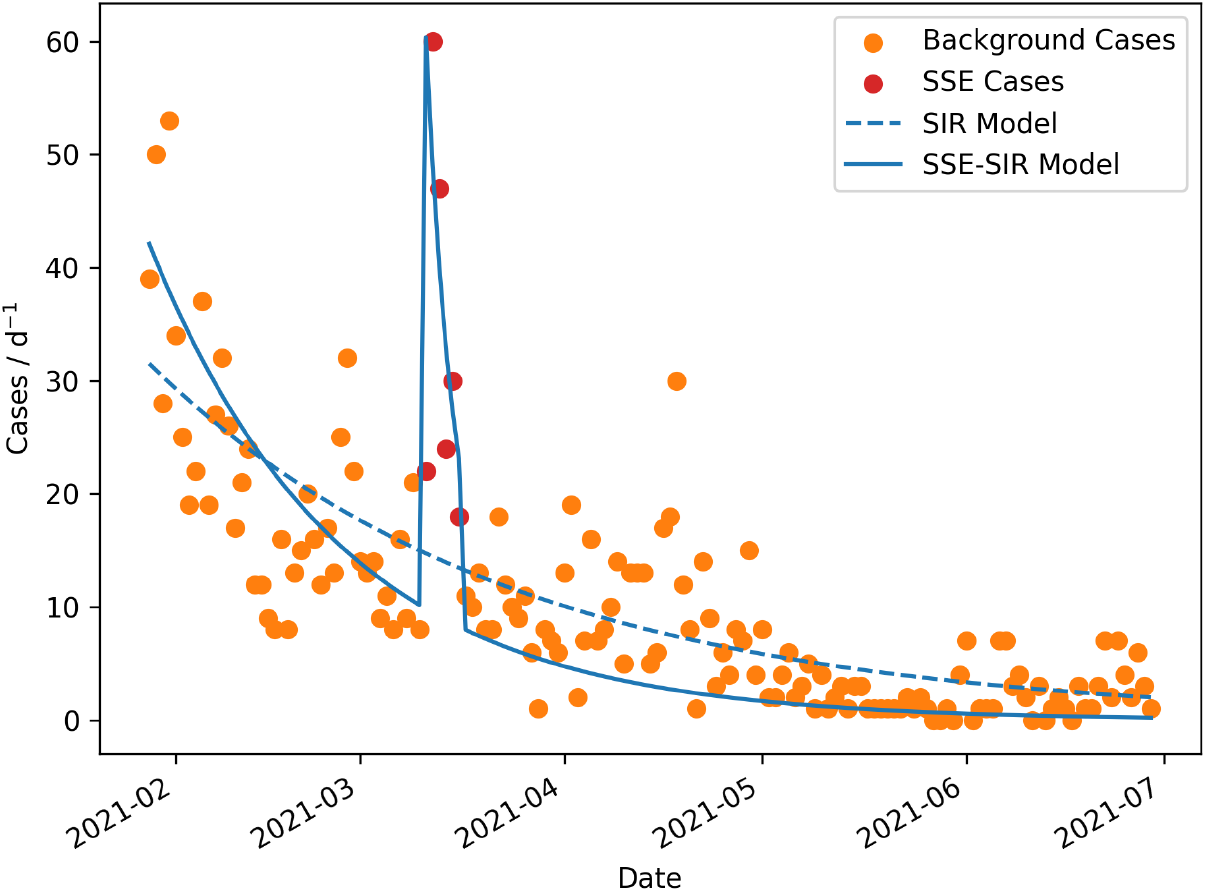
The traditional SIR model (1) - (3) (dashed blue line) and the SSE SIR model (4) - (6) fitted to the reported daily cases (orange dots) and the SSE (red dots) in Hong Kong.

The SSE has a duration of *t*_*S*_ = 6 d and a maximum amplitude of *a* = 50 d^−1^. Weibull function parameter estimates of *τ* = 2.2 and *k* = 0.68, yielding a model that significantly differs from an exponential model, which would have *k* = 1. The total sum of modelled residuals is 147 cases, with 102 cases that could be directly linked to the SSE [53].

We now focus on a case study with both predictable and unpredictable time variations in transmission rates in Gütersloh (Germany). The time period spans from the beginning of the first wave in Germany early March 2020 until the beginning of the Autumn wave in August 2020. This period includes six predictable changes in NPIs and the unpredictable SSE in June 2020. We used piecewise constant transmission rates that adjust with each introduction or relaxation of NPIs [40] (Fig. 4). Not accounting for the SSE and including the SSE-related case numbers together with the background cases into the parameter estimation of the SIR model caused the modelled case numbers to increase over the sixth NPI period, which poorly matches the observed cases before and after the SSE, while also not matching the SSE itself at all. Treating the SSE cases as outliers and parameterising the SSE-SIR model results in a much better fit. The actual SSE is reproduced reasonably well with the SSE model (7), with the exception of two days at the very end of the event. Additionally, the cases before and after the SSE are modelled much better. Looking at the transmission rate estimates, we see that the rate decreases from *β*_6_ = 0.19 d^−1^ to 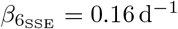 when explicitly modelling the SSE. This is a relative difference of 16%, with 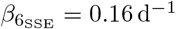 surely being much closer to the actual transmission rate averaged over the whole of the Gütersloh district with a population of 360,000, as is required by homogeneity assumption of SIR models. We only compare the statistics of the model fits for the 6th NPI period, as the models are equal until then. When accounting for the SSE, the MSE improves from 1.9 to 0.64 and the SSPB improves from -0.61 to -0.27. Most strikingly, the Pearson correlation coefficient even improves from an anticorrelation of -0.40 to 0.75.

**Fig 4.**
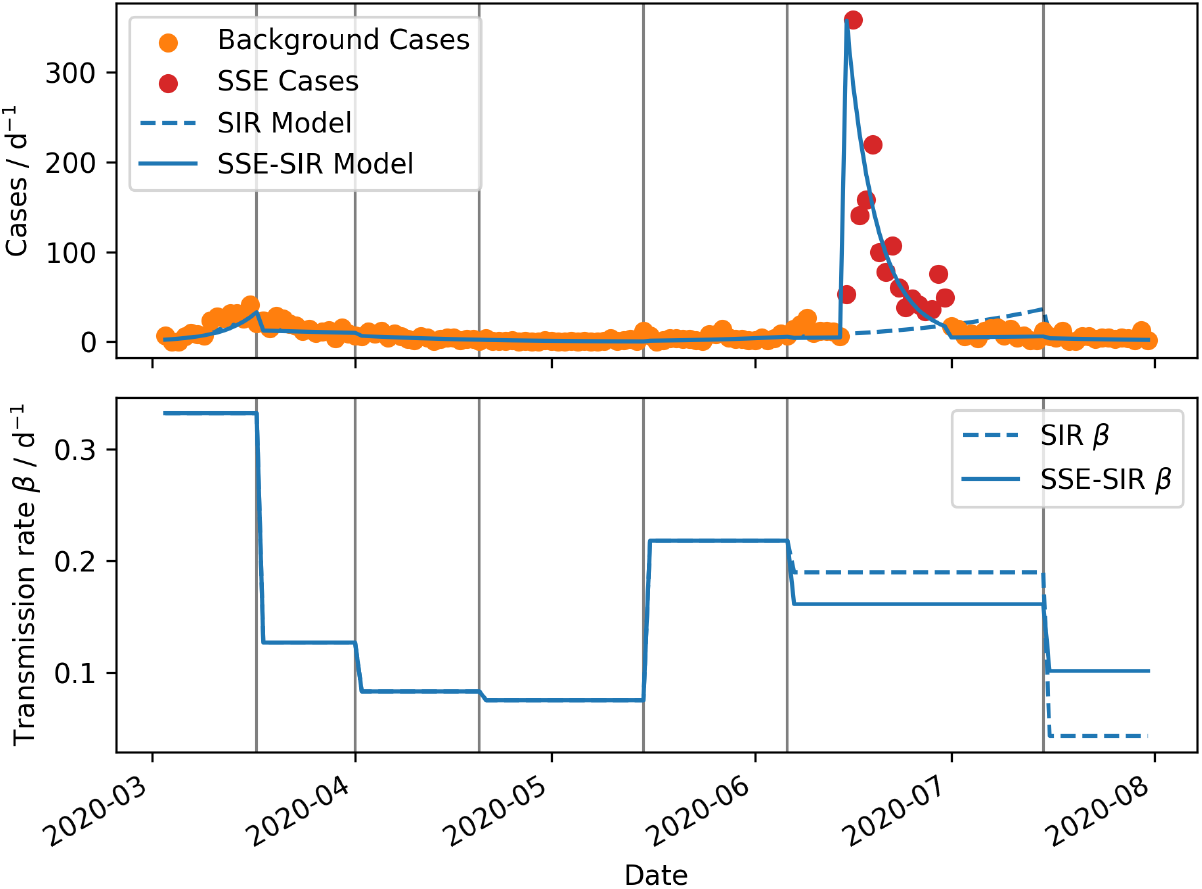
Upper panel: The orange dots indicate the daily cases reported to the RKI for the district of Gütersloh, Germany. The red dots show the case numbers during the identified SSE. The dashed blue curve shows the result of the SIR model without SSEs - (3) with parameters calibrated towards the RKI data. The solid blue curve shows the result of the SSE-SIR model (5) - (6) with parameters calibrated towards the RKI data without the SSE, and (7) with parameters calibrated towards the identified SSE. The horizontal black lines show the dates when NPIs where implemented or relaxed. Lower panel: The dashed blue curve shows the estimated transmission rate *β* of the SIR model. The solid blue curve shows the estimated transmission rate *β* of the SSE-SIR model.

Looking at the transmission rates of the final NPI period of this time series, we see another problem of the SIR model caused by the SSE. To match the cases here, the model has to decrease *β* to nearly 0, causing a much larger difference between the transmission rates estimated with and without the SSE component. The exact estimates of both model estimates are *β*_7_ = 0.044 d^−1^ and 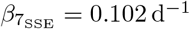, with a relative difference of 233%. The parameterisation of the SSE model (7) yields a duration of *t*_*S*_ = 16 d and a maximum amplitude of *a* = 353 d^−1^. The estimated Weibull parameters are *τ* = 3.2 and *k* = 1.1, which suggests that the SSE decay model can be simplified to an exponential function as the shape parameter k is close to 1. The total sum of residuals is 1485 cases, with 1413 infections directly linked to the SSE and 353 secondary infections identified [54].

## Discussion

In this work, we presented novel methods and models to deal with heterogeneities arising from SSEs in the context of SIR-type compartment models for infectious diseases. We developed a modified SIR-type compartment model based on the model introduced by Schüler et al. 2021 [40]. The original model took a first step to include heterogeneities in transmission rates induced by NPI changes through policy decisions. This is achieved by using piecewise constant transmission rates. To this model (1) - (3), we added a new pathway between the *S*usceptible and the *R*ecovered compartments (4) - (6), which transfers the residuals of the detected SSEs. The SSEs themselves are modelled with a Weibull function (7), which has a total of four parameters, the duration *t*_S_, the maximum amplitude *a*, and two shape parameters *τ* and *k*. Using the compound PPP (8), we created a fully generative model of SSEs that allowed us to simulate synthetic data under the SSE-SIR model with which we could test our approach. To identify and quantify potential SSEs, we used the uLOF outlier detection method applied to time series of daily cases.

With this modular setup, we created an ensemble of 1000 realisations of the PPP (8) and modelled each one with both, the Schüler et al. SIR (1) - (3) and the new SSE-SIR model (4) - (6). We then compared the results between the two models based on three key metrics, the MSE, SSPB, and Pearson’s correlation coefficient. The SSE-SIR model demonstrated significant improvements over the SIR model across all metrics. Maybe even more important, the resulting estimated transmission rates are much more meaningful, because they do not have to account for the local SSE transmissions, which can cause extremely skewed averaged transmission rates. Consequently, the underlying epidemiological dynamics can be modelled more accurately, while we simultaneously characterise and learn about SSEs, specifically their frequency, magnitude, duration, and shape.

Rather than relying on complex models and assumptions to simulate heterogeneities such as SSEs, employing outlier detection methods alongside simple SIR-type models offers the substantial advantage of requiring minimal data. As soon as case count data are available, the methods presented here can be readily applied.

The individual parts of this framework work together in a modular way, meaning that individual components such as the outlier detection method, the Weibull model of SSEs (7), or the compound PPP (8), can easily be swapped out to tailor the model to specific situations. It is worth mentioning that the specific parameters of the generative model, like the lognormal distribution of the case number’s error term were also chosen to be as simple as possible

We chose to illustrate our approach with the uLOF outlier detection method due to its simplicity, interpretability, and easy accessibility. However, its mediocre performance represents a clear area where our approach can be improved, although this will likely come at the cost of more complicated outlier detection methods. Indeed outlier detection is an active area of statistical research [45], and these efforts have notably expanded since the advent of machine learning methods. An obvious next step is therefore to find a more specialised, better performing SSE identification method. One of the most promising current research topic in the field of outlier detection is using deep learning methods [55], specifically applied to multivariate time series [56]. We see potential in using multivariate time series in the context of SSE detection due to the enormous amounts of data already constantly being collected. A promising and relatively straightforward idea to improve our approach is to incorporate neighbouring districts into the outlier detection step. A limitation of the current univariate outlier detection method is that relatively small SSEs may be obscured by background noise and thus not get flagged unless the outlier detection is made overly sensitive, which in turn would increase the false positive rate. One potential solution to this problem would be to compare the focal district with adjacent districts via multivariate outlier detection. Specifically, an absence of the same peak in neighbouring time series can potentially increase our confidence that such a relatively small local peak is indeed an SSE. Conversely, very large SSEs can generate secondary smaller peaks in neighbouring districts (for example the Gütersloh event), and incorporating those spatially correlated signals would improve the robustness of the SSE detection.

In this study, we only introduced a new pathway from the *S*usceptible to the *R*ecovered compartments, with the assumption that especially large SSEs quickly get noticed by authorities, who can then react with NPIs like quarantining, which makes secondary cases unlikely. However, by introducing a new parameter *ν* ∈ [0, 1] we can control the transmissions going from SSEs to either the infectious compartment, with probability *ν*, or to the recovered compartment, with probability 1 − *ν*. This generalisation can also be applied to different compartments in more complex SIR-type models.

## Data Availability

All data produced in the present work are currently available upon reasonable request to the authors and will be available online once the manuscript is published by a journal.

https://github.com/owid/covid-19-data/blob/master/public/data/README.md

https://pypi.org/project/pycocap/

## References

1. Lloyd-Smith JO, Schreiber SJ, Kopp PE, Getz WM. Superspreading and the effect of individual variation on disease emergence. Nature. 2005;438(7066):355–359. doi:10.1038/nature04153.

2. Leo Y, Chen M, Heng B, Lee C. Severe acute respiratory syndrome–Singapore, 2003. MMWR Morbidity and mortality weekly report. 2003;52(18):405–411.

3. Shen Z, Ning F, Zhou W, He X, Lin C, Chin DP, et al. Superspreading SARS Events, Beijing, 2003. Emerging Infectious Diseases. 2004;10(2):256–260. doi:10.3201/eid1002.030732.

4. Dye C, Gay N. Modeling the SARS Epidemic. Science. 2003;300(5627):1884–1885. doi:10.1126/science.1086925.

5. Lipsitch M, Cohen T, Cooper B, Robins JM, Ma S, James L, et al. Transmission Dynamics and Control of Severe Acute Respiratory Syndrome. Science. 2003;300(5627):1966–1970. doi:10.1126/science.1086616.

6. Riley S, Fraser C, Donnelly CA, Ghani AC, Abu-Raddad LJ, Hedley AJ, et al. Transmission Dynamics of the Etiological Agent of SARS in Hong Kong: Impact of Public Health Interventions. Science. 2003;300(5627):1961–1966. doi:10.1126/science.1086478.

7. Bauch CT, Lloyd-Smith JO, Coffee MP, Galvani AP. Dynamically Modeling SARS and Other Newly Emerging Respiratory Illnesses: Past, Present, and Future. Epidemiology. 2005;16(6):791–801. doi:10.1097/01.ede.0000181633.80269.4c.

8. Lemieux JE, Siddle KJ, Shaw BM, Loreth C, Schaffner SF, Gladden-Young A, et al. Phylogenetic analysis of SARS-CoV-2 in the Boston area highlights the role of recurrent importation and superspreading events. Epidemiology; 2020.

9. Tabata S, Imai K, Kawano S, Ikeda M, Kodama T, Miyoshi K, et al. Clinical characteristics of COVID-19 in 104 people with SARS-CoV-2 infection on the Diamond Princess cruise ship a retrospective analysis. The Lancet Infectious Diseases. 2020;20(9):1043–1050. doi:10.1016/S1473-3099(20)30482-5.

10. Wei WE, Li Z, Chiew CJ, Yong SE, Toh MP, Lee VJ. Presymptomatic Transmission of SARS-CoV-2 — Singapore, January 23–March 16, 2020. MMWR Morbidity and Mortality Weekly Report. 2020;69(14):411–415. doi:10.15585/mmwr.mm6914e1.

11. McMichael TM, Currie DW, Clark S, Pogosjans S, Kay M, Schwartz NG, et al. Epidemiology of Covid-19 in a Long-Term Care Facility in King County, Washington. New England Journal of Medicine. 2020;382(21):2005–2011. doi:10.1056/NEJMoa2005412.

12. Baggett TP, Keyes H, Sporn N, Gaeta JM. Prevalence of SARS-CoV-2 Infection in Residents of a Large Homeless Shelter in Boston. JAMA. 2020;323(21):2191. doi:10.1001/jama.2020.6887.

13. Frieden TR, Lee CT. Identifying and Interrupting Superspreading Events—Implications for Control of Severe Acute Respiratory Syndrome Coronavirus 2. Emerging Infectious Diseases. 2020;26(6):1059–1066. doi:10.3201/eid2606.200495.

14. Guenther T, Czech-Sioli M, Indenbirken D, Robitailles A, Tenhaken P, Exner M, et al. Investigation of a superspreading event preceding the largest meat processing plant-related SARS-Coronavirus 2 outbreak in Germany. SSRN Electronic Journal. 2020;doi:10.2139/ssrn.3654517.

15. Kermack WO, McKendrick AG. A Contribution to the Mathematical Theory of Epidemics. Proceedings of the royal society of london Series A. 1927;115(772):700–721.

16. Diekmann O, Heesterbeek JAP, Metz JAJ. On the definition and the computation of the basic reproduction ratio R 0 in models for infectious diseases in heterogeneous populations. Journal of Mathematical Biology. 1990;28(4). doi:10.1007/bf00178324.

17. Anderson RM, May RM. Infectious diseases of humans: dynamics and control. Oxford: Oxford Univ. Press; 1992.

18. Hethcote HW. The Mathematics of Infectious Diseases. SIAM Review. 2000;42(4):599–653. doi:10.1137/s0036144500371907.

19. Keeling MJ, Rohani P. Modeling infectious diseases in humans and animals. Princeton: Princeton University Press; 2008.

20. Brauer F. In: Brauer F, van den Driessche P, Wu J, editors. Compartmental Models in Epidemiology. Berlin, Heidelberg: Springer Berlin Heidelberg; 2008. p. 19–79. Available from: 10.1007/978-3-540-78911-6_2.

21. Rvachev LA, Longini IM. A mathematical model for the global spread of influenza. Mathematical Biosciences. 1985;75(1):3–22. doi:10.1016/0025-5564(85)90064-1.

22. Bjørnstad ON, Finkenstädt BF, Grenfell BT. DYNAMICS OF MEASLES EPIDEMICS: ESTIMATING SCALING OF TRANSMISSION RATES USING A TIME SERIES SIR MODEL. Ecological Monographs. 2002;72(2):169–184. doi:10.1890/0012-9615(2002)072[0169:domees]2.0.co;2.

23. Granich RM, Gilks CF, Dye C, De Cock KM, Williams BG. Universal voluntary HIV testing with immediate antiretroviral therapy as a strategy for elimination of HIV transmission: a mathematical model. The Lancet. 2009;373(9657):48–57. doi:10.1016/s0140-6736(08)61697-9.

24. Chen SC, Hsieh MH. Modeling the transmission dynamics of dengue fever: Implications of temperature effects. Science of The Total Environment. 2012;431:385–391. doi:10.1016/j.scitotenv.2012.05.012.

25. Rachah A, Torres DFM. Mathematical Modelling, Simulation, and Optimal Control of the 2014 Ebola Outbreak in West Africa. Discrete Dynamics in Nature and Society. 2015;2015:1–9. doi:10.1155/2015/842792.

26. Adiga A, Dubhashi D, Lewis B, Marathe M, Venkatramanan S, Vullikanti A. Mathematical Models for COVID-19 Pandemic: A Comparative Analysis. Journal of the Indian Institute of Science. 2020;100(4):793–807. doi:10.1007/s41745-020-00200-6.

27. Jose SA, Raja R, Omede BI, Agarwal RP, Alzabut J, Cao J, et al. Mathematical modeling on co-infection: transmission dynamics of Zika virus and Dengue fever. Nonlinear Dynamics. 2023;111(5):4879–4914. doi:10.1007/s11071-022-08063-5.

28. Flahault A, Vergu E, Coudeville L, Grais RF. Strategies for containing a global influenza pandemic. Vaccine. 2006;24(44-46):6751–6755. doi:10.1016/j.vaccine.2006.05.079.

29. Choi S, Jung E. Optimal Tuberculosis Prevention and Control Strategy from a Mathematical Model Based on Real Data. Bulletin of Mathematical Biology. 2014;76(7):1566–1589. doi:10.1007/s11538-014-9962-6.

30. Knerer G, Currie CSM, Brailsford SC. Impact of combined vector-control and vaccination strategies on transmission dynamics of dengue fever: a model-based analysis. Health Care Management Science. 2015;18(2):205–217. doi:10.1007/s10729-013-9263-x.

31. Flaxman S, Mishra S, Gandy A, Unwin HJT, Mellan TA, Coupland H, et al. Estimating the effects of non-pharmaceutical interventions on COVID-19 in Europe. Nature. 2020;584(7820):257–261. doi:10.1038/s41586-020-2405-7.

32. Kissler SM, Tedijanto C, Goldstein E, Grad YH, Lipsitch M. Projecting the transmission dynamics of SARS-CoV-2 through the postpandemic period. Science. 2020;368(6493):860–868. doi:10.1126/science.abb5793.

33. Agbomola JO, Loyinmi AC. Modelling the impact of some control strategies on the transmission dynamics of Ebola virus in human-bat population: An optimal control analysis. Heliyon. 2022;8(12):e12121. doi:10.1016/j.heliyon.2022.e12121.

34. Calabrese JM, Schüler L, Fu X, Gawel E, Zozmann H, Bumberger J, et al. A novel, scenario-based approach to comparing non-pharmaceutical intervention strategies across nations. Journal of The Royal Society Interface. 2024;21(218). doi:10.1098/rsif.2024.0301.

35. Naaly BZ, Marijani T, Isdory A, Ndendya JZ. Mathematical modeling of the effects of vector control, treatment and mass awareness on the transmission dynamics of dengue fever. Computer Methods and Programs in Biomedicine Update. 2024;6:100159. doi:10.1016/j.cmpbup.2024.100159.

36. Brauer F, Castillo-Chavez C, Feng Z. Mathematical Models in Epidemiology. Texts in Applied Mathematics. New York, NY: Springer New York; 2019. Available from: https://link.springer.com/10.1007/978-1-4939-9828-9.

37. Kermack WO, McKendrick AG. Contributions to the mathematical theory of epidemics. II. —The problem of endemicity. Proceedings of the Royal Society of London Series A, Containing Papers of a Mathematical and Physical Character. 1932;138(834):55–83. doi:10.1098/rspa.1932.0171.

38. Ferguson NM, Cummings DAT, Cauchemez S, Fraser C, Riley S, Meeyai A, et al. Strategies for containing an emerging influenza pandemic in Southeast Asia. Nature. 2005;437(7056):209–214. doi:10.1038/nature04017.

39. Libotte GB, dos Anjos L, Almeida RC, Malta SMC, Silva RS. Framework for enhancing the estimation of model parameters for data with a high level of uncertainty. Epidemiology; 2020. Available from: http://medrxiv.org/lookup/doi/10.1101/2020.12.17.20248389.

40. Schüler L, Calabrese JM, Attinger S. Data driven high resolution modeling and spatial analyses of the COVID-19 pandemic in Germany. PLOS ONE. 2021;16(8):e0254660. doi:10.1371/journal.pone.0254660.

41. Zozmann H, Schüler L, Fu X, Gawel E. Autonomous and policy-induced behavior change during the COVID-19 pandemic: Towards understanding and modeling the interplay of behavioral adaptation. PLOS ONE. 2024;19(5):e0296145. doi:10.1371/journal.pone.0296145.

42. Iglewicz B, Hoaglin DC. How to detect and handle outliers. No. 16 in The ASQC basic references in quality control: statistical techniques. Milwaukee, Wis: ASQC Quality Press; 1993.

43. Hodge V, Austin J. A Survey of Outlier Detection Methodologies. Artificial Intelligence Review. 2004;22(2):85–126. doi:10.1023/b:aire.0000045502.10941.a9.

44. Blázquez-García A, Conde A, Mori U, Lozano JA. A Review on Outlier/Anomaly Detection in Time Series Data. ACM Computing Surveys. 2022;54(3):1–33. doi:10.1145/3444690.

45. Boniol P, Paparrizos J, Palpanas T. An Interactive Dive into Time-Series Anomaly Detection. In: 2024 IEEE 40th International Conference on Data Engineering (ICDE). Utrecht, Netherlands: IEEE; 2024. p. 5382–5386. Available from: https://ieeexplore.ieee.org/document/10597392/.

46. Peirce CS. The Numerical Measure of the Success of Predictions. Science. 1884;ns-4(93):453–454. doi:10.1126/science.ns-4.93.453.b.

47. Youden WJ. Index for rating diagnostic tests. Cancer. 1950;3(1):32–35. doi:10.1002/1097-0142(1950)3:1¡32::aid-cncr2820030106¿3.0.co;2-3.

48. Breunig MM, Kriegel HP, Ng RT, Sander J. LOF: identifying density-based local outliers. ACM SIGMOD Record. 2000;29(2):93–104. doi:10.1145/335191.335388.

49. Schmidt L, Schäfer D, Geller J, Lünenschloss P, Palm B, Rinke K, et al. System for automated Quality Control (SaQC) to enable traceable and reproducible data streams in environmental science. Environmental Modelling Software. 2023;169:05809. doi:10.1016/j.envsoft.2023.105809.

50. Schäfer D, Palm B, Lünenschloß P, Schmidt L, Schnicke T, Bumberger J. System for automated Quality Control-SaQC; 2024. Available from: https://zenodo.org/doi/10.5281/zenodo.5888547.

51. Schüler L. pyCOCAP; 2024. Available from:https://zenodo.org/doi/10.5281/zenodo.16563488.

52. Habershon S, Nenoff K, Kraemer G, Schüler L, Zozmann H, Calabrese JM, et al. The spatiotemporal dynamics of COVID-19 in Europe: time-series clustering maps 5 distinct trajectories to spatial patterns. Population Health Metrics. 2025;23(1):44. doi:10.1186/s12963-025-00405-w.

53. Chu DK, Gu H, Chang LD, Cheuk SS, Gurung S, Krishnan P, et al. SARS-CoV-2 Superspread in Fitness Center, Hong Kong, China, March 2021. Emerging Infectious Diseases;27(8):2230. doi:10.3201/eid2708.210833.

54. Landtag Nordrhein-Westfalen Plenarprotokoll 17/94; 2020. Available from: https://www.landtag.nrw.de/portal/WWW/dokumentenarchiv/Dokument? Id=MMP17/94%7C9%7C36.

55. Zamanzadeh Darban Z, Webb GI, Pan S, Aggarwal C, Salehi M. Deep Learning for Time Series Anomaly Detection: A Survey. ACM Computing Surveys. 2025;57(1):1–42. doi:10.1145/3691338.

56. Yahya MA, Moya AR, Ventura S. Deep learning for multivariate time series anomaly detection: an evaluation of reconstruction-based methods. Artificial Intelligence Review. 2025;58(12):400. doi:10.1007/s10462-025-11401-9.

