## Supplementary material for "Using outlier detection methods to incorporate highly heterogeneous infection rates into compartment models"

**1** Dept. of Computational Hydrosystems, UFZ – Helmholtz Centre for Environmental Research, Leipzig, Germany
**2** Research Data Management, UFZ – Helmholtz Centre for Environmental Research, Leipzig, Germany
**3** Dept. Monitoring and Exploration Technologies, UFZ – Helmholtz Centre for Environmental Research, Leipzig, Germany
**4** Center for Advanced Systems Understanding (CASUS), Görlitz, Germany
**5** German Centre for Integrative Biodiversity Research (iDiv) Halle-Jena-Leipzig, Puschstraße 4, 04103 Leipzig, Germany
**6** Helmholtz-Zentrum Dresden Rossendorf (HZDR), Dresden, Germany
**7** Dept. of Ecological Modelling, UFZ – Helmholtz Centre for Environmental Research, Leipzig, Germany

*

### Supporting information

#### uLOF outlier detection

The LOF assigns a score to data points in multi-dimensional datasets to indicate their anomaly status based on local data distribution variations. It effectively identifies outliers in datasets with varying local densities by evaluating the local neighbourhood of each point.

The locality of a point $p$ is defined by its $k$-nearest neighbours $N_k(p)$. The distance is defined as

$$d_k(p,q) = \max(d(p,q), \max_{s \in N_k(q)}(d(q,s))) \tag{1}$$

and the average distance $\hat{D}_k(p)$ to these neighbours is calculated as

$$D_k(p) = \frac{1}{k} \sum_{q \in N_k(p)} d_k(p,q). \tag{2}$$

This average distance inversely correlates with local density: higher $D_k$ indicates lower density. The LOF score for $p$ is now derived by comparing $D_k(p)$ to the distances of its neighbours:

$$\mathrm{LOF}(p) = \frac{1}{k} \sum_{q \in N_k} \frac{D_k(p)}{D_k(q)}. \tag{3}$$

To apply LOF to univariate time series data $X = (X_t; n \in \mathbb{N})$, the values must be embedded in a space that reflects both magnitude and temporal distance. This is achieved by constructing a dataset $XY$:

$$XY = \{(X(n), n * \hat{\mu}|\Delta X|) | n \in \mathbb{N}\}, \tag{4}$$

where $\hat{\mu}$ is an aggregation function (e.g., the median $\tilde{\mu}$), and $\Delta$ is the discrete derivative operator. The univariate Local Outlier Factor (uLOF) is then derived as:

$$uLOF(X) = LOF(XY). \tag{5}$$

This approach allows for the computation of anomaly scores that reflect the expected differences between consecutive values in $X$.

To classify a value $X(n)$ as normal or anomalous, a binary flag $f_n$ is assigned based on a threshold $t$:

$$f_n(t) := \begin{cases} 1, & uLOF(X)(n) > t \\ 0, & uLOF(X)(n) \leq t. \end{cases} \tag{6}$$

A single threshold can be used across the dataset, although dynamic thresholds may be considered for varying local distributions. The threshold $t$ can be tuned based on visual inspection of scores or through optimisation against a flagged dataset.

If outliers are not present in the tuning dataset, $t$ can be set to the supremum of the normal data scores with a small offset. If outliers may be present, $t$ can be derived by removing the upper $\alpha$-quantile from the scores and setting $t$ as the supremum of the remainder with an offset.

The implementation of the uLOF in the SaQC Python package [1, 2], we use for this study, is defined by the function signature:

```
flagUniLOF(k=20, thresh=1.5, p=1, min_offset=0)
```

Parameters include `k` (number of neighbours), `thresh` (cut-off threshold), `p` (norm type), and `min_offset` (to prevent overflagging (false positives), e.g. of small increases in low variance regimes or small context windows). This allows for the incorporation of domain knowledge into the anomaly detection process.

The parameters can either be manually adjusted, with interpretable parameters proving to be especially helpful or they can be optimised. However, common gradient based algorithms will not work here, because one of the parameters is an integer, creating large areas in the parameter space with a gradient of zero. Luckily, there are only two parameters with reasonably tight bounds, making even a brute force algorithm computationally feasible, with no noticeable computational time.

#### uLOF applied to generative model

We have evaluated the uLOF outlier detection method by applying it to the 1000 realisations of the generative compound Poisson point process (8), of which 25 realisations are shown together with the detected outliers in a matrix of 5 by 5 plots (Fig. 1). We note that in this particular case, only a single data point is often identified as a false positive and indeed, it sits on top the first wave's peak, being significantly larger than the rest of the case numbers. Furthermore, looking at plot $(3, 1)$, we see that relatively small SSEs during a highly dynamic situation like the first wave cannot be detected with the uLOF method. However, in many other situations, the algorithm can successfully detect the SSEs, with the exception of the tails of SSEs with longer durations. Applying Youden's J statistic [3, 4], also known as the true skill statistic, to the whole ensemble results in a mean detection score of $\bar{J} = 0.6$, a standard deviation of $\sigma_J = 0.2$, a minimum score of $J_{\min} = 0.1$, and a maximum score of $J_{\max} = 1$.

### References


1. Schmidt L, Schäfer D, Geller J, Lünenschloss P, Palm B, Rinke K, et al. System for automated Quality Control (SaQC) to enable traceable and reproducible data

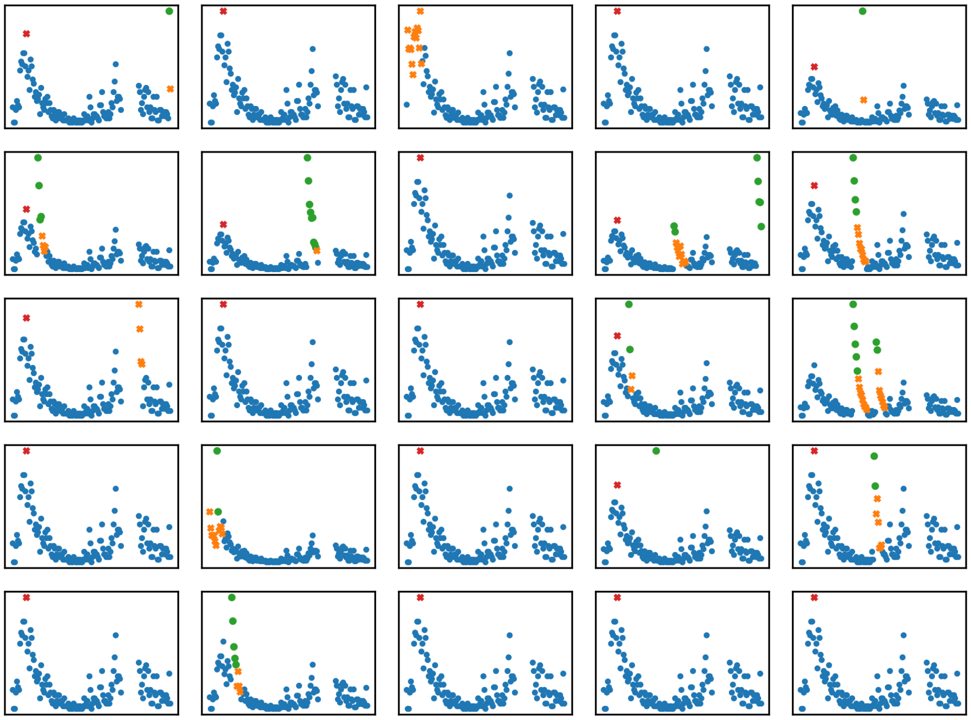

**Fig 1.** The uLOF outlier detection method applied to 25 Realizations of the compound Poisson point process (8). The correctly identified SSEs are shown as green dots, the false negative SSEs are shown as orange crosses and the false positive cases are shown as red crosses. For the sake of clarity, the axes labels were omitted, for reference see Fig. 4.

streams in environmental science. Environmental Modelling  Software. 2023;169:105809. doi:https://doi.org/10.1016/j.envsoft.2023.105809.

2. Schäfer D, Palm B, Lünenschloß P, Schmidt L, Schnicke T, Bumberger J. System for automated Quality Control - SaQC; 2024. Available from: https://zenodo.org/doi/10.5281/zenodo.5888547.

3. Peirce CS. The Numerical Measure of the Success of Predictions. Science. 1884;ns-4(93):453–454. doi:10.1126/science.ns-4.93.453.b.

4. Youden WJ. Index for rating diagnostic tests. Cancer. 1950;3(1):32–35. doi:10.1002/1097-0142(1950)3:1¡32::aid-cncr2820030106¿3.0.co;2-3.
